# Predictors of white matter hyperintensities in the elderly Congolese population

**DOI:** 10.1101/2024.09.03.24313022

**Authors:** Emile Omba Yohe, Alvaro Alonso, Daniel L. Drane, Saranya Sundaram Patel, Megan Schwinne, Emmanuel Epenge, Guy Gikelekele, Esambo Herve, Immaculee Kavugho, Nathan Tshengele, Samuel Mampunza, Lelo Mananga, Liping Zhao, Deqiang Qiu, Anthony Stringer, Amit M Saindane, Jean Ikanga

**Author notes:** Co-Corresponding authors: please send general questions about the study to Emile Omba Yohe and Dr. Jean Ikanga. Jean N. Ikanga, Ph.D. Department of Rehabilitation Medicine Emory University 1441 Clifton Rd NE Atlanta, GA 30322, USA (404)727-2844 (office phone).

## Abstract

**Background:** White matter hyperintensities (WMHs) are strongly linked to cardiovascular risk factors and other health conditions such as Alzheimer’s disease. However, there is a dearth of research on this topic in low-income countries and underserved populations, especially in the Democratic Republic of Congo (DRC) where the population is aging rapidly with increasing cardiovascular risk factors and dementia-related diseases. This study evaluates health factors associated with WMH in the elderly Sub-Saharan Africa (SSA), specifically Congolese adults.

**Methods:** In a cross-sectional study of 77 people from the DRC, participants underwent neuroimaging to analyze WMH volume and completed clinical evaluation, laboratory-based blood exams, self-reported questionnaires, and interviews. A simple linear regression model was conducted to test the association between WMH and potential predictors (neurological status, age, sex, hypertension, diabetes, tobacco abuse, stroke, high cholesterol, cardiovascular medication, and alcohol abuse). Stepwise selection and backward elimination analyses were performed to obtain the final model. Finally, a multiple linear regression model was conducted to assess the association between WMH and variables retained in the final model (neurological status, sex, and age).

**Results:** Of the 77 individuals, 47 (61%) had dementia, 40 (52.6%) were males, and the mean age was 73 years (± 8.0 years standard deviation). In simple linear regression models, WMH was significantly associated with dementia (expβ1=1.75, 95% CI=1.14 – 2.71, p-value=0.01) though it had a weak association with age (expβ1=1.03, 95% CI=1.00 – 1.05, p-value=0.05) and sex (male) (expβ1=0.66, 95% CI=0.43 – 1.01, p-value=0.05). In multiple linear regression models, WMH was statistically significantly associated with dementia (expβ1=1.97, 95% CI=1.31 – 2.95, p-value =0.001), male sex (expβ2=0.54, 95% CI=0.36 – 0.80, p-value=0.003), and age (expβ3=1.03, 95% CI=1.00 – 1.06, p-value=0.03). However, WMH was not significantly associated with common cardiovascular risk factors, such as high blood pressure, diabetes, tobacco use, obesity, and high cholesterol levels.

**Conclusion:** WMH is significantly associated with neurological status, sex, and age in the Congolese population. Understanding these predictors may improve our ability to diagnose, assess, and develop preventative treatments for white matter disease in SSA/DRC populations, where neuroimaging is difficult to obtain.

## INTRODUCTION

White matter hyperintensities (WMHs), or lesions, are a marker of change to the connective nerve fibers of the brain, potentially contributing to cognitive decline, balance, and mobility issues.^1,2^ WMHs are present in approximately 20-50% of the general population in midlife, increasing to more than 90% with advanced age^2,3^ and can be related to normal aging changes or neurodegenerative diseases, such as Alzheimer’s disease (AD).^4,5^ AD is the leading cause of dementia in individuals aged 65 or older, with vascular dementia being the second most common.^6,7^ AD is a significant global burden present in approximately 40-50 million people and it is predicted to be over 150 million by 2050.^5,8,9^ WMHs are found in more than 89% of patients with AD and are typically more severe than WMHs seen in older adults without dementia.^10,11,12^ The observed patterns of WMHs can reflect the advanced phase of the disease, as significant and anatomically congruent correlations between WMHs and regional gray matter atrophy have been seen in patients with AD.^11^

The prevalence of white matter disease increases with age, with rates of more than 90% in older adults.^2^ In vascular dementia, research has found that WMHs are strongly linked to cardiovascular disease risk factors (e.g., smoking, hypertension, diabetes, dyslipidemia, physical inactivity, being overweight, and obesity) as a consequence of cSVD.^13,14,15^ Furthermore, researchers have demonstrated that the association between WMHs and early signs of cognitive decline is detectable many years before the emergence of clinical symptoms of AD and related dementia (ADRD); therefore, WMHs can be used as a surrogate biomarker of cognitive decline and risk of dementia.^16^ Interestingly, the prevalence of WMHs is greater in the female sex, leading to a higher incidence and prevalence of dementia in women than in men in older adults.^17^

There has been substantial research documenting the association between cardiovascular risk factors, AD, and WMHs in high-income countries (HIC) and populations of European ancestry. However, few studies have investigated the increasing risk of WMHs in low- and middle-income countries (LMIC).^18^ The predicted increase in dementia-related diseases like WMHs is more susceptible in LMIC, especially in East Asia and Sub-Saharan Africa (SSA), where 70% of the population will be with dementia in 2040.^19,20^ However, there is limited research on WMHs in the DRC where the population is aging rapidly with high prevalence rates of cardiovascular risk factors and suspected ADRD. Thus, further research is needed in the DRC to understand health factors associated with WMHs to determine whether age, CV risk factors, and ADRD are associated with WMHs in this population. As WMHs are slowly progressive, early detection will provide the opportunity for early intervention to reduce or delay the occurrence of dementia-related outcomes, such as memory problems, balance issues, and mood changes in the population. This study evaluates factors associated with WMHs in a cohort from the elderly Congolese population. We hypothesized that WMHs will be associated with AD-related dementia, CV risk factors and disease, and age.

## METHODS

### Study design and population

This cross-sectional study was conducted from 2019 to 2022 in Kinshasa, DRC. Initially, 1432 people were selected through a community-based recruitment procedure from door-to-door in the city, clinics, hospitals, churches, and older adult associations. Inclusion criteria included 50 years of age or older, diagnosis of major neurocognitive disorder or normal cognition according to DSM-5, having a collateral informant, and being fluent in French or Lingala. Participants were excluded from this study if they had a history of schizophrenia, neurological, or other medical conditions potentially affecting the central nervous system (CNS). Due to the lack of clear cutoff values for AD biomarkers in the SSA to clinically confirm the diagnosis of probable AD, we used two screening measures with high sensitivity and specificity for identifying individuals with dementia in Western cohorts, the Community Screening Instruments for Dementia (CSID) and Alzheimer’s Questionnaire (AQ).^21, 22^ The AQ distinguishes between those with AD from healthy controls. The CSID Questionnaire has been extensively used in many international and SSA dementia studies, including studies in Nigeria, Uganda, and South Africa.^22^ Based on cognitive and functional deficits for Diagnostic and Statistical Manual of Mental Disorders, Fifth Edition, Text Revision (DSM-5-TR) diagnostic criteria, we used Brazzaville cut-offs of CSID, the closest city from Kinshasa, to classify participants. Similar to our prior study, participants were classified using CSID and AQ scores (Figure 1), which yielded 4 groups: major neurocognitive disorder/dementia, mild neurocognitive disorder (MND), subjective cognitive impairment, and healthy control (HC), i.e., normal cognition. For the AQ, only the total score was used, based on 27 possible points and a cutoff score of 13 or more points suggestive of dementia. Written informed consent was obtained from all participants before undergoing any study procedures approved by the Ethical Committee and Institutional Review Boards of the University of Kinshasa. All participants were financially compensated for their time.^21^

**Figure 1:**
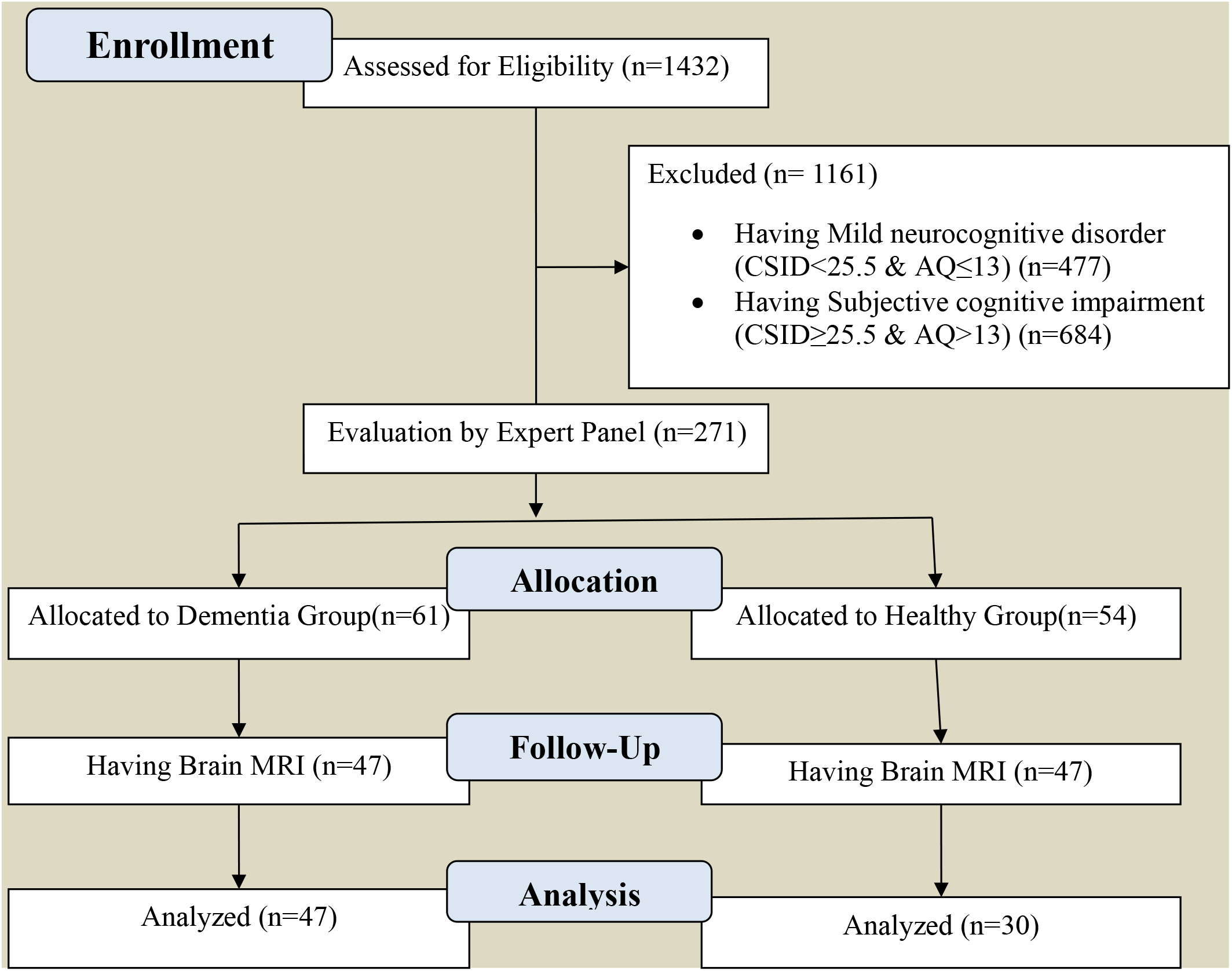
Flow Diagram of Participant Recruitment

A panel consisting of a neurologist, psychiatrist, and neuropsychologist reviewed screening tests, clinical interviews, and neurological examination of subjects. 61 individuals were confirmed with a diagnosis of major neurocognitive disorders and 54 were deemed HCs. HCs were matched based on age, sex, and education levels before the MRI procedure. Brain MRI was then performed, resulting in the final sample of 77 subjects, 47 with dementia and 30 HCs.

### Measures

Demographic, socioeconomic, and medical history (e.g., stroke, tobacco use, alcohol use, and cardiovascular medication) information were obtained through participants’ self-reported questionnaires and interviews. Participants were categorized into four different age groups: 50- 64, 65-74, 75-84, and 85+ and four different education groups: primary school (1-6 years), secondary school (7-12 years), some or completion of university (13-17 years), and beyond university (18+ years).^21^

Three blood pressure measurements were conducted by a medical resident at complete rest. A participant was considered to have hypertension if their systolic blood pressure was greater than 139 mmHg and diastolic was greater than 89 mmHg. Participants were considered to have diabetes mellitus type 2 if their blood sugar had an HbA1C level ≥ 6.5 % or having a fasting blood glucose ≥ 7.0 mmol/l. The cholesterol level was detected by blood test. Participants were considered to have high cholesterol if their total cholesterol is above 200 mg/dl.

### Neuroimaging Parameters

Each participant was scanned using a 1.5 Tesla MRI unit scanner (Siemens, Magneton Sonata) at HJ Hospitals in Kinshasa, DRC, using the same standardized imaging acquisition protocol based on the Alzheimer’s Disease Research Center (ADRC) protocol of Emory University.^22^ It consisted of sagittal volumetric T1-weighted (MP-RAGE), coronal T2-weighted, axial diffusion-weighted, T2-weighted, and T2-FLAIRE sequences. High-resolution structural images were obtained using a T1-weighted MP-RAGE Sequence with the following parameters: Repetition Time (TR) = 2200 ms; minimum full Echo time (TE) = 1000 ms; Flip angle = 8^0^; Field of view (FOV) = 250 mm; acquisition matrix= 192 X 184, yielding a voxel size of approximately 1.25 mm X 1.25mm X 1.2 mm.

### Quantification of WMHs

To identify and extract WMH volume, brain segmentation was performed by experts at Emory University. An experienced subspecialty-certified neuroradiologist reviewed images to ensure that automated processes resulted in reasonable data (AMS). WMHs were graded according to the age-related white matter changes (ARWMC) scales.^22^ The number of chronic brain parenchymal micro hemorrhages was recorded, and the lobar volume loss pattern of the brain was assessed. Regional brain volume for both cortical and sub-cortical brain regions was calculated. Finally, the presence or absence of any additional abnormalities was noted, and participants with neuroimaging evidence indicated an etiology other than probable AD (e.g., the presence of a brain tumor), were excluded from the analysis.

The Fazekas scale for white matter lesions was used to quantify the amount of white matter T2 hyperintense lesions,^22^ which is mainly used to describe white matter disease severity. It divides the white matter into periventricular and deep white matter. Each region is graded based on the size and confluence of lesions. For this analysis, we used the deep white matter (DWM) classification, which is crucial in assessing patients with possible dementia and the component that is usually reported in clinical research (e.g., Fazekas grade 2).^22^:

- 0 = absent
- 1 = punctate foci
- 2 = beginning confluence
- 3 = large confluence areas.

### Statistical Analysis

All statistical analyses were completed using SAS version 9.4 statistical software. Descriptive statistics (e.g., frequencies, percentages, means, and standard errors) were generated for both the overall sample and between neurological status (Table 1). WMHs volumes were log-transformed for normality. A univariable analysis in a simple linear regression model was conducted to test the association between WMHs (dependent variable) and each predictor (primary independent variable). Neurological status, age, age decade, sex, hypertension, diabetes, tobacco abuse, stroke, high cholesterol, cardiovascular medication, and alcohol abuse predictors were considered predictors. A multivariable analysis in a multiple linear regression model was conducted to assess the association of WMHs and all the predictors retained in the final model. Multivariable analysis started with a stepwise selection using a p-value cut-off of 0.20 for variable entry and removal. Then, backward elimination was performed to remove insignificant variables at the 0.05 level to obtain the final model. The predictors that were retained in the final model were neurological status, sex, and age. The results were expressed as exp (beta) coefficients with corresponding 95% confidence intervals. All statistical tests were two-sided; p-values < 0.05 were considered statistically significant.

**Table 1:**
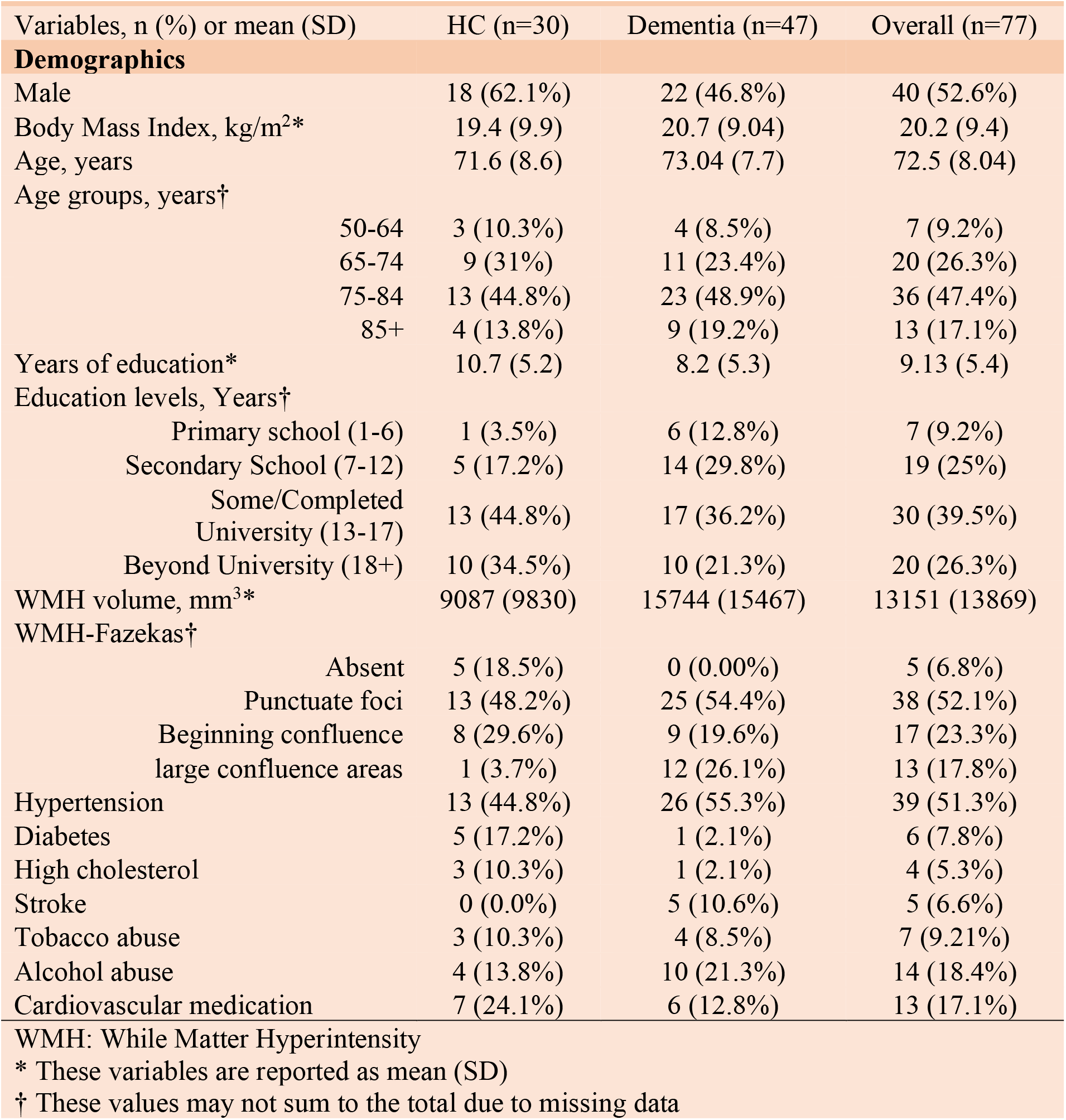
Descriptive Characteristics of the sample population, stratified by Neurological Status.

| Variables, n (%) or mean (SD) | HC (n=30) | Dementia (n=47) | Overall (n=77) |
| --- | --- | --- | --- |
| **Demographics** |  |  |  |
| Male | 18 (62.1%) | 22 (46.8%) | 40 (52.6%) |
| Body Mass Index, kg/m<sup>2</sup>* | 19.4 (9.9) | 20.7 (9.04) | 20.2 (9.4) |
| Age, years | 71.6 (8.6) | 73.04 (7.7) | 72.5 (8.04) |
| Age groups, years† |  |  |  |
| 50-64 | 3 (10.3%) | 4 (8.5%) | 7 (9.2%) |
| 65-74 | 9 (31%) | 11 (23.4%) | 20 (26.3%) |
| 75-84 | 13 (44.8%) | 23 (48.9%) | 36 (47.4%) |
| 85+ | 4 (13.8%) | 9 (19.2%) | 13 (17.1%) |
| Years of education* | 10.7 (5.2) | 8.2 (5.3) | 9.13 (5.4) |
| Education levels, Years† |  |  |  |
| Primary school (1-6) | 1 (3.5%) | 6 (12.8%) | 7 (9.2%) |
| Secondary School (7-12) | 5 (17.2%) | 14 (29.8%) | 19 (25%) |
| Some/Completed University (13-17) | 13 (44.8%) | 17 (36.2%) | 30 (39.5%) |
| Beyond University (18+) | 10 (34.5%) | 10 (21.3%) | 20 (26.3%) |
| WMH volume, mm<sup>3</sup>* | 9087 (9830) | 15744 (15467) | 13151 (13869) |
| WMH-Fazekas† |  |  |  |
| Absent | 5 (18.5%) | 0 (0.00%) | 5 (6.8%) |
| Punctuate foci | 13 (48.2%) | 25 (54.4%) | 38 (52.1%) |
| Beginning confluence | 8 (29.6%) | 9 (19.6%) | 17 (23.3%) |
| large confluence areas | 1 (3.7%) | 12 (26.1%) | 13 (17.8%) |
| Hypertension | 13 (44.8%) | 26 (55.3%) | 39 (51.3%) |
| Diabetes | 5 (17.2%) | 1 (2.1%) | 6 (7.8%) |
| High cholesterol | 3 (10.3%) | 1 (2.1%) | 4 (5.3%) |
| Stroke | 0 (0.0%) | 5 (10.6%) | 5 (6.6%) |
| Tobacco abuse | 3 (10.3%) | 4 (8.5%) | 7 (9.21%) |
| Alcohol abuse | 4 (13.8%) | 10 (21.3%) | 14 (18.4%) |
| Cardiovascular medication | 7 (24.1%) | 6 (12.8%) | 13 (17.1%) |
| WMH: While Matter Hyperintensity |  |  |  |
| * These variables are reported as mean (SD) |  |  |  |
| † These values may not sum to the total due to missing data |  |  |  |

## RESULTS

### Descriptive characteristics of the sample population

Participants were evenly distributed between males (52.6%) and females (47.4%). The mean age was 73 years and participants’ age groups, education levels, and body mass index showed no significant differences between dementia and control groups. Regarding WMHs, the sample mean WMHs volume was 13,151 mm^3^, with dementia cases having a higher mean WMH volume (15,745 mm^3^) than HC (9,088 mm^3^). A little more than half of the participants (51.3%) had hypertension, with a higher prevalence among dementia compared to HC cases (55.3% and 44.8%, respectively). Additionally, more dementia cases (11%) reported a history of stroke compared to the healthy control cases (0%).

### Association between WMHs and predictors

The association between WMHs and its predictors was performed using the log-transformed value of WMHs volume (Figure 2) in simple and multiple linear regression models.

**Figure 2:**
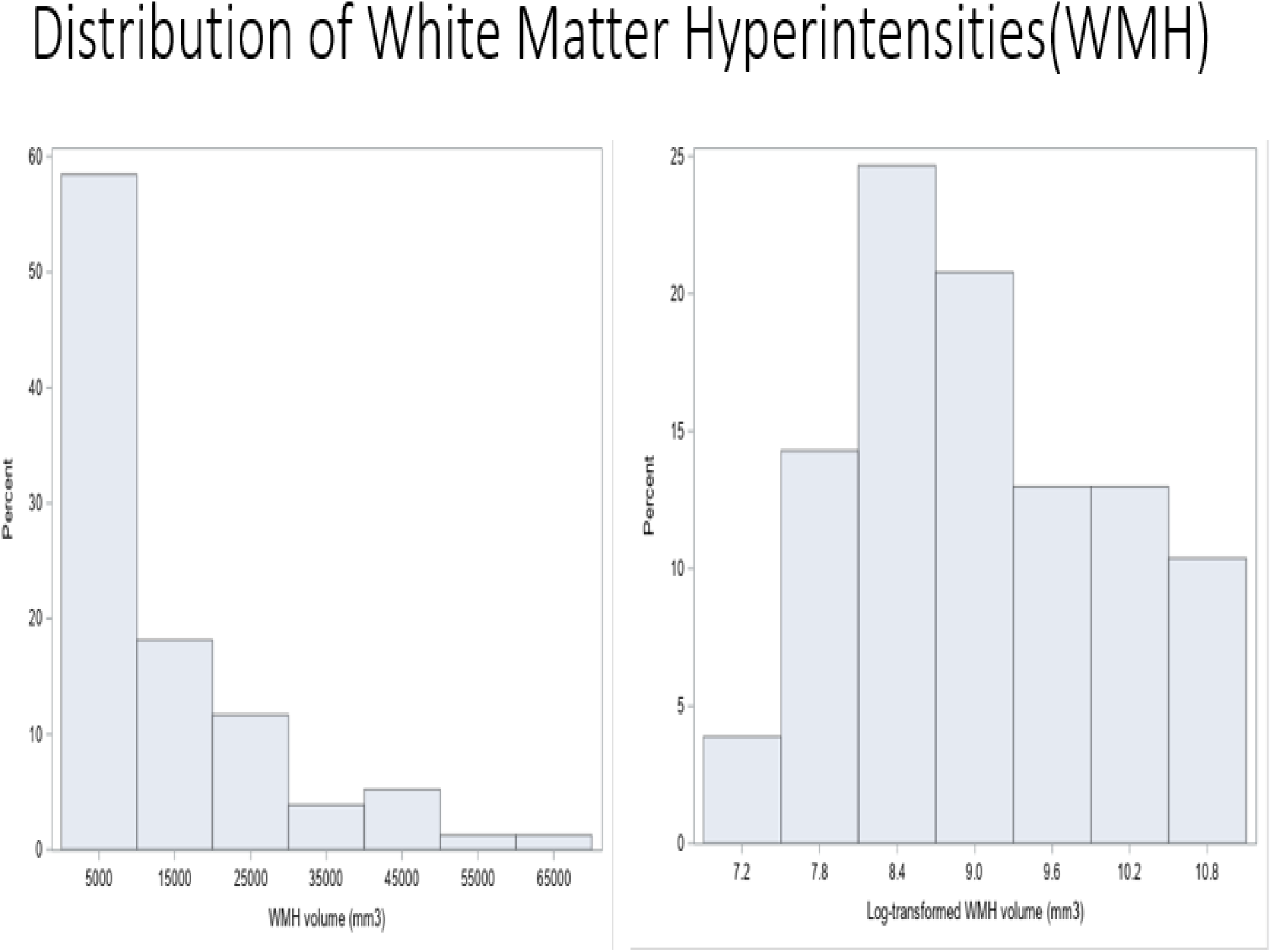

### Association between WMHs and each predictor

In a simple linear regression model, WMH was significantly associated with neurological status (dementia) (expβ1=1.75, 95% CI=1.14 – 2.71, p-value=0.01). Participants with dementia had a 75% increase in WMH volume compared to HC. WMH had a statistically weak association with age (expβ1=1.03, 95% CI=1.00 – 1.05, p-value=0.05) and sex (male) (expβ1=0.66, 95% CI=0.43 – 1.01, p-value=0.05). Male participants had a WMH volume 34% lower than female participants, and when participants’ age increases by 1 year, it is associated with a 3% increase in WMH volume. There were no statistically significant associations between WMH and other predictors, such as age decade (expβ1=1.21, 95% CI=0.94 – 1.56, p-value=0.12), diabetes (expβ1=1.84, 95% CI=0.82 – 4.10, p-value=0.13), alcohol abuse (expβ1=0.67, 95% CI=0.38 – 1.16, p-value=0.15), tobacco abuse (expβ1=1.66, 95% CI=0.78 – 3.50, p-value=0.18), stroke (expβ1=0.72, 95% CI=0.32 – 1.58, p-value=0.40), high cholesterol (expβ1=1.42, 95% CI=0.53 – 3.79, p-value=0.48), hypertension (expβ1=0.85, 95% CI=0.56 – 1.35, p-value=0.54), or cardiovascular medication (expβ1=0.93, 95% CI=0.52 – 1.67, p-value=0.81) [Table 2].

**Table 2:** Associations between WMHs and predictors.

| Variable | Exp($\beta$) | 95%CI | p-value |
| --- | --- | --- | --- |
| Neurological status | 1.75 | 1.14-2.71 | 0.01 |
| Age (total years) * | 1.03 | 1.00-1.05 | 0.05 |
| Age decade | 1.21 | 0.94-1.56 | 0.14 |
| Sex (male) | 0.66 | 0.43-1.01 | 0.05 |
| Hypertension | 0.87 | 0.56-1.35 | 0.54 |
| Diabetes | 1.84 | 0.82-4.10 | 0.13 |
| Tobacco abuse | 1.66 | 0.78-3.5 | 0.18 |
| Stroke | 0.72 | 0.32-1.58 | 0.4 |
| High Cholesterol | 1.42 | 0.53-3.79 | 0.48 |
| Cardiovascular medication | 0.93 | 0.52-1.67 | 0.81 |
| Alcohol abuse | 0.67 | 0.38-1.16 | 0.15 |

WMH: White Matter Hyperintensity.
\* These variables are reported as continuous

### Association between WMH and final model variables

The summary of the stepwise selection process showed that four variables were selected: neurological status (expβ1=1.95, 95% CI=1.30 – 2.93, p-value=0.001), sex (Male) (expβ2=0.53, 95% CI=0.36 – 0.80, p-value=0.003), age (expβ3=1.03, 95% CI=1.00 – 1.05, p-value=0.03), and tobacco abuse (expβ4=1.59, 95% CI=0.81 – 3.12, p-value=0.17) [Table 3].

**Table 3:** Summary of stepwis selection.

| Variable | Exp($\beta$) | 95%CI | p-value |
| --- | --- | --- | --- |
| Neurological status | 1.95 | 1.30-2.93 | 0.001 |
| Sex (Male) | 0.53 | 0.36-0.80 | 0.003 |
| Age, per year* | 1.03 | 1.00-1.05 | 0.03 |
| Tobacco abuse | 1.59 | 0.81-3.12 | 0.17 |

\* These variables are reported as continuous.

Stepwise selection uses a p-value cut-off of 0.20 for variable entry and removal. Our results are presented as the exponentiated beta value of each predictor considered in a multiple linear regression model. The result corresponds to the relative increase or decrease in WMH volume associated with a specific variable.

Then, after the backward elimination process, only three variables were retained in our final model: neurological status (expβ1=1.97, 95% CI=1.31 – 2.95, p-value=0.001), sex (male) (expβ2=0.54, 95% CI=0.36 – 0.80, p-value=0.003), and age (expβ3=1.03, 95% CI=1.00 – 1.06, p-value=0.03) [Table 4].

**Table 4:**
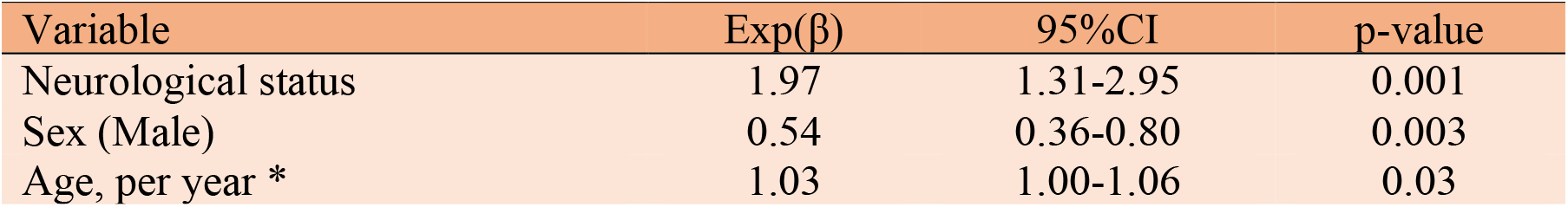
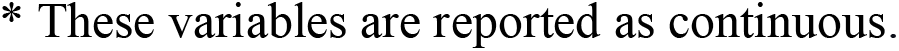
Summary of backward elimination/final model.

| Table 4: Summary of backward elimination/final model |  |  |  |
| --- | --- | --- | --- |
| Variable | Exp($\beta$) | 95%CI | p-value |
| Neurological status | 1.97 | 1.31-2.95 | 0.001 |
| Sex (Male) | 0.54 | 0.36-0.80 | 0.003 |
| Age, per year * | 1.03 | 1.00-1.06 | 0.03 |

\* These variables are reported as continuous.

The multivariable analysis showed a statistically significant association between WMH and all three variables in the final model. Neurological status (dementia) was significantly associated with WMH when controlling sex and age. Participants with dementia had two times higher (97% increase) volume of WMH than the HC, controlling for sex and age. Sex (male) was significantly less associated with WMH, when adjusted for neurological status and age. Male participants had a 46% decrease in WMH volume compared to females, controlling for neurological status and age. Age was significantly associated with WMH, controlling for neurological status and sex. The increase in 1 year of age was associated with a 3% increase in participants’ WMH volume, controlling for neurological status and sex [Table 4].

Backward elimination was performed to remove insignificant variables at the 0.05 level to obtain the final model. Our results are presented as the exponentiated beta value of each predictor considered in the final model of a multiple linear regression model. The result corresponds to the relative increase or decrease in WMH volume associated with a specific variable.

## DISCUSSION

This study evaluated health factors associated with WMHs in the elderly community-based sample from Kinshasa, DRC. We hypothesized that WMHs will be associated with AD-related dementia, CV risk factors and diseases, and age. For the unadjusted analysis, we found a significant association between WMH and neurological status and a weak association between WMH and the sex and age of the participants. Participants with dementia had a 75% increase in WMH volume compared to HC. Male participants had a 34% decrease in WMH volume than female, and the increase in age by 1 year is associated with a 3% increase in WMH volume. In the adjusted analysis, dementia status, older age, and female sex were significantly associated with a larger volume of WMHs independently of each other. Participants with dementia had almost twice (97% increase) the volume of WMHs than the HC, controlling for sex and age.

Male participants had a 46 % decrease in WMH volume compared to female, when adjusted for neurological status and age. The increase in 1 year of age was associated with a 3% increase in participants’ WMHs volume, controlling for neurological status and sex. Our results were meaningfully similar to published studies of populations in HIC, where previous researchers suggest that WMHs are associated with an increased risk of cognitive dysfunction and are found in most patients with AD-related dementia.^12, 23^ Additionally, researchers suggest that white matter diseases start at midlife, and their prevalence increases as people get older, with more than 90% of WMH cases seen in older adults.^2^ Moreover, previous studies showed an association between WMH and sex, as the female population had a higher incidence and prevalence of white matter diseases than the male population.^17^ These previous results align with our findings that WMHs were significantly associated with neurological status (dementia), sex, and age of participants both in an univariable and multivariable analysis.

WMHs are confirmatory imaging diagnosis of changes in the brain through MRI, which most patients are not able to afford the cost in the DRC. Clinically, the presence of WMHs on MRI is associated with cognitive decline, balance issues, and mood change (depression).^2^ Our findings can be used clinically as helpful scientific evidence that will enable physicians and healthcare professionals to consider WMHs as a differential diagnostic for patients with dementia-related problems and gait disturbance even without having the MRI result. In the population level, our findings will be essential in implementing public health programs that target individual behavioral change to prevent WMHs and stop white matter diseases from getting worse, which will be relevant in decreasing the burden of dementia (disability and loss of life, social isolation, discrimination, stigmatization, and death) in the Congolese population. White matter disease is common in the general population. They can be caused by numerous factors, such as cardiovascular risks and disease, AD-related dementia, age, and sex.

Understanding the association between WMHs and its predictors is crucial in implementing early diagnosis, prevention programs, and treatment. This study found that WMHs are significantly associated with neurological status (dementia), sex (females), and age (older adults) in the Congolese population. Knowing these predictors of WMH in the Congolese population could be a useful prevention tool for white matter disease in SSA countries, especially in DRC, where brain MRI diagnosis is difficult to obtain.

Limitations include a relatively small sample size (77 participants) which can subsequently reduce the power of the study and limited the detection of differences that could have been clinically and significantly relevant to discriminate the two groups. Thus, future studies should replicate these findings with larger sample sizes. Second, the screening measures used (CSID and AQ) were not yet validated in the SSA/DRC. Third, this study included 2 categories of participants which are suspected dementia and healthy controls. Participants who were seen in between the spectrum (e.g., MCI, subjective memory complaints) were excluded, leaving only the extremes of the dementia spectrum. Future studies should include all the 4 groups (healthy controls, MCI, subjective memory complaint and dementia). Other limitations include lack of normative data for the fluid biomarkers used here in the studied population, relatively small sample size for a biomarker study, and no replication cohort. Cardiovascular risk factors also appeared relatively low in this sample. Future studies should aim to replicate our findings along the AD pathology continuum, tau pathology, neurofilament light (NfL), as well as use other plasma biomarkers (e.g., glial fibrillar acidic protein [GFAP]), as they may provider greater insight into the progression of cerebral amyloid and tau pathology and cognitive decline in SSA populations. A major caveat is that amyloid positive was determined with plasma Aβ42/40 by Simoa, which is far from being the best biomarker for amyloid positivity. The gold standard is amyloid brain PET. As this is the first WMH-related study to be done in DRC, more studies with larger sample sizes are needed in which the association between WMH and its predictors is examined from the same population for better comparison and further validation of other predictors that were not significantly associated with WMH in our study, like cardiovascular.

## Data Availability

All data produced in the present study are available upon reasonable request to the authors

## ACKNOWLEDGMENTS

The authors acknowledged the assistance and expertise from the Medical Center of Kinshasa and HJ Hospitals who have assisted our study with magnetic resonance imaging used in the study and other materials.

## FUNDING

This study was supported by grants AARG-19-61778701 and P30AG066511-02S1 from the National Institute on Aging.

Dr. Alonso was supported by NIH/NHLBI grant K24HL148521 and NIH/NIA grant P30AG06651.

Dr. Drane and the neuroimaging analysis efforts were funded by the NIH/NINDS through grant R01 NS088748.

MS was supported by the National Center for Advancing Translational Sciences of the National Institutes of Health under Award Number UL1TR002378. The content is solely the responsibility of the authors and does not necessarily represent the official views of the National Institutes of Health.

## CONFLICT OF INTEREST

The authors have no conflict of interest to report.

## DATA AVAILABILITY

The data supporting the findings of this study are available on request from the corresponding author. The data are not publicly available due to privacy or ethical reasons.

## Notes

### Competing Interest Statement

The authors have declared no competing interest.

### Author Declarations

The procedures were approved by the Ethics Committee/Institutional Review Boards of the University of Kinshasa.

